# Symptom Associations with Delayed Gastric Emptying Vary by Disease Phenotype

**DOI:** 10.64898/2026.07.14.26358018

**Authors:** Jayden Kumar, Chris Varghese, I-Hsuan Huang, Stefan Calder, Gabriel Schamberg, Nicky Dachs, Sam Simmonds, Daphne Foong, Christopher N Andrews, Armen A Gharibans, Jan Tack, Greg O’Grady

**Affiliations:** Department of Surgery, University of Auckland, Auckland, New Zealand; Alimetry Ltd, Auckland, New Zealand; Translational Research Center in Gastro-intestinal Disorders, KU Leuven, Leuven, Belgium

**Author notes:** **Corresponding author**: Professor Greg O’Grady Department of Surgery University of Auckland New Zealand. **Disclosures:** G.O. and A.A.G. hold intellectual property and grants in gastric electrophysiology and are Directors of University of Auckland spin-out company Alimetry Ltd; C.V., S.C., G.S., N.D., S.S., D.F., and C.N.A. are members of Alimetry Ltd. J.K., I.H., and J.T. have no relevant conflicts of interest to declare.

**Keywords:** Gastroparesis, functional dyspepsia, Body Surface Gastric Mapping

## Abstract

**Background & Aims:** The association between upper gastrointestinal (GI) symptoms and delayed gastric emptying time (GET) is debated. This study utilized Body Surface Gastric Mapping (BSGM) and the ‘Auckland Classification’ BSGM phenotyping scheme to investigate whether symptom associations with GET vary across different mechanistic phenotypes.

**Methods:** A pooled analysis was performed on two prospective datasets of 194 patients with chronic upper GI symptoms. Participants underwent simultaneous BSGM (Gastric Alimetry) and gastric emptying testing (breath test or scintigraphy). Validated time-of-test symptom profiling was recorded at 15-minute intervals using 0-10 Likert-type scales. Patients were classified: ‘Spectral Abnormal’ (abnormal electrophysiology), ‘Sensorimotor’ (symptoms correlate with gastric electrical amplitude), ‘Continuous’ (symptoms uncorrelated), and ‘NA’ (no phenotype).

**Results:** In the analyzed cohort, 24% had delayed GET. Overall, delayed GET was associated with a higher total symptom burden (15.6 delayed vs. 8.8 normal, p=0.006; r=0.30, p<0.001), specifically postprandial fullness (p=0.004) and early satiation (p=0.015). However, associations varied significantly by phenotype. In the ‘Sensorimotor’ phenotype, nausea was associated with delayed GET (4.9 vs. 1.0, p=0.027; r=0.59, p=0.009), while total symptom burden was 23.3 vs. 8.4 (p=0.135). Conversely, patients with ‘Continuous’ or ‘Spectral Abnormal’ phenotypes, or who were unclassified, showed no symptom associations.

**Conclusions:** Delayed GET is weakly associated with an increased burden of upper GI symptoms. However, gastroduodenal disorders are heterogeneous, such that phenotyping reveals this association to be exclusive to patients with a ‘Sensorimotor’ phenotype. These results could improve targeting of therapies that address gastric emptying.

## Introduction

Chronic gastroduodenal symptoms, such as nausea, epigastric pain, and postprandial distress, impose a substantial global health burden and are a frequent driver of healthcare utilization, affecting >10% of the global population ^1,2^. Defining, diagnosing and treating the disorders underlying these symptoms is challenging owing in part to their overlapping symptom, testing and diagnostic profiles ^3,4^.

Traditionally, the clinical management of these patients has been informed by the assessment of gastric emptying, with the presence of delayed gastric emptying aiding in the differentiation of gastroparesis from functional dyspepsia ^5^. However, the clinical utility of this distinction is debated ^6–8^. One area of focus has been the inconsistent associations between symptoms and gastric emptying ^6,9^. Given the heterogeneity of these disorders and the discordant findings across existing literature, a clear relationship between symptoms and gastric emptying has remained elusive ^6,10–12^. Methodological limitations and incomplete disease classification may be contributory. For example, traditional studies, sometimes coupled with sub-optimal gastric emptying testing protocols, have often measured symptoms using retrospective questionnaires, which are prone to recall bias and fail to capture the dynamic nature of postprandial symptoms ^10,13,14^. In addition, it is plausible that certain subgroups within the gastroparesis and functional dyspepsia spectrum may show stronger symptom associations with delayed gastric emptying than others ^15^.

This study aimed to address these gaps by utilizing time-of-test symptom profiling aided by a validated App ^16^, as well as by employing patient phenotyping with the newly-established ‘Auckland Classification’ scheme [17]. This mechanistic classification employs Body Surface Gastric Mapping (BSGM) and digital symptom profiling to classify chronic gastroduodenal disorders. The hypothesis was that symptoms would be associated with delayed gastric emptying for some patient subgroups (i.e. the ‘Sensorimotor’ phenotype, in which symptoms are correlated with gastric electrophysiological amplitude), but not associated for others (i.e. the ‘Continuous’ phenotype, in which symptoms are continuous and uncorrelated with gastric electrophysiological amplitude).

## Methods

This study was a pooled analysis of two prospectively-collected datasets comprising patients with chronic gastroduodenal symptoms (N=214). In one dataset (n=143), patients were recruited as part of an observational cohort study conducted in Leuven, Belgium, investigating simultaneous BSGM and Gastric Emptying Breath Testing (GEBT). The study protocol was approved by the Ethics Committee Research UZ Leuven (S65541) ^17^. In the second dataset (n=71), patients were recruited as part of a study conducted in Auckland, New Zealand and Western Sydney, Australia, investigating simultaneous BSGM and Gastric Emptying Scintigraphy (GES). The study protocol was approved by the Auckland Health Research Ethics Committee (AHREC123) and the Western Sydney University Human Research Ethics Committee (H13541) ^18^. All participants provided informed consent.

### Inclusion and Exclusion Criteria

The pooled analysis included patients aged ≥18 years, presenting with chronic gastroduodenal symptoms. A prerequisite for inclusion was an upper gastrointestinal endoscopy to exclude alternative pathologies. Furthermore, patients were excluded if they had a known structural gastrointestinal disease or a history of prior gastric surgery. Specific exclusion criteria related to Gastric Alimetry included allergies to adhesives or compromised epigastric skin.

In addition, from the initial pooled cohort of 214 patients (143 GEBT + 71 GES), the study population was further refined as follows:

1. General Analysis Cohort (n=194): twenty patients were excluded due to rapid gastric emptying (t_1/2_ <45 minutes (GEBT)), inadequate meal completion (<50%), or insufficient GEBT data. This cohort was used for the initial analysis of symptom associations with gastric emptying time (GET), without patient phenotyping.
2. ‘Auckland Classification’ Cohort (n=155): a further 39 patients were excluded due to specific exclusion criteria related to Gastric Alimetry BSGM testing. This included: those with a BMI >35 (higher BMIs warrant cautious interpretation due to signal attenuation), and those with BSGM testing with a high percentage artifact (>50%) coupled with a low Gastric Alimetry Rhythm Index (GA-RI <0.25)^19^.

### Test Methodologies

Gastric Alimetry was measured simultaneously with standardized ^13^C octanoic acid solid GEBT or standardized 99^m^Tc GES. All patients fasted overnight prior to testing, and were asked to stop medications known to affect gastric emptying, including, but not limited to, opioids, prokinetics, anticholinergics, calcium channel blockers, for at least 2 days prior to testing. Further, patients were asked to avoid caffeine, nicotine and cannabis on the day of testing. For GEBT, the test meal was either a pancake with 180mL of water (11.2g fat, 31.7g carbohydrate, 8.4g protein; 261 kcal total) or an egg with two slices of white toast and 180mL of water (9.4g fat, 34.0g carbohydrate, 11.5g protein; 268kcal total) ^17^, whilst for GES, the test meal was a standardized low-fat egg meal (255kcal total) ^13^. For GEBT, breath samples were collected prior to the test meal and at 15-minute intervals during the 4-hour postprandial period, whilst for GES, scintigraphy imaging was performed at 1-hour intervals during the 4-hour postprandial period.

For both datasets, Body Surface Gastric Mapping (BSGM) was performed using the Gastric Alimetry system (Alimetry, Auckland, New Zealand) as per standardized methods ^20^. The system includes a stretchable array of electrodes (8x8 electrodes; 20-mm interelectrode spacing; 196cm^2^ area), a wearable Reader, and a validated App with time-of-test symptom logging ^21,22^. BSGM recordings were performed over a preprandial fasting period of 30 minutes, during the test meal, and over a postprandial period of at least four hours. Patients sat in a reclined chair and were asked to limit movement, talking and sleeping.

Time-of-test symptom profiling occurred using the validated Gastric Alimetry app, with the aid of pictograms ^16^. For continuous symptoms (nausea, excessive fullness, bloating, upper gut pain, stomach burn, and heartburn), profiling occurred at 15-minute intervals (30-minutes preprandial to 4-hours postprandial) using an 11-point Likert-type scale ranging from 0 (no symptoms) to 10 (worst imaginable extent of symptoms). For discrete symptoms (reflux, belching, and vomiting), symptoms were recorded on a per event basis. Early satiation was measured immediately after meal completion. Symptom data was combined algorithmically to create a total symptom burden according to standardized methods ^16^.

### Data Classification

For categorical comparisons, delayed GET was defined as t_1/2_ >109 minutes (GEBT) or >10% retention at 4 hours (GES) according to existing clinical standards ^13,17^. The remainder of patients were classified as having a normal GET.

Patients were phenotyped according to the recently-developed ‘Auckland Classification’ scheme for chronic gastroduodenal disorders ^23^. This scheme utilizes both BSGM spectral data and time-of-testing symptom data to classify individuals into six distinct categories (individuals can be classified into one category, no categories, or more than one category) ^18,24^:

- ‘Spectral Abnormal’:

○ ‘Dysrhythmic’: defined by the presence of an unstable gastric slow wave rhythm (GA-RI <0.25) ^19^, considered associated with gastric neuromuscular dysfunction;
○ ‘High Frequency’: defined by gastric slow wave frequency >3.35;
○ ‘Low Meal Response’: defined by a low meal response ratio ^17^, reflecting weak postprandial activity;
- ‘Sensorimotor’: defined as gastric amplitude correlation with symptoms >0.5;
- ‘Continuous’: defined by normal spectral metrics with meal- and amplitude-independent symptom profiles;
- ‘Delayed Symptom Onset’: defined as late onset of symptoms, often associated with small intestinal symptom contributions ^25^.

Those with ‘Dysrhythmic’, ‘High Frequency’ and ‘Low Meal Response’ phenotypes were grouped as ‘Spectral Abnormal’ due to a lack of individual statistical power. If patients were classified into more than one ‘Auckland Classification’ category, they were included in both categories independently. Furthermore, an insufficient number of patients were classified into the ‘Delayed Symptom Onset’ category, and thus these individuals were grouped with those who did not fit a distinct category as ‘Not Applicable’.

### Statistical Analysis

All statistical analyses were performed using R version 4.5.2 (R Foundation for Statistical Computing, Vienna, Austria). Continuous variables were reported as medians due to the non-normal distribution of data, having been defined as such by Shapiro-Wilk testing. Categorical variables were reported as frequencies or percentages. Comparisons of symptom burden between normal and delayed GET groups were performed using the Mann-Whitney U test. Correlations between symptom burden and GET were performed using Spearman’s rank correlation coefficient. Chi-squared and Fisher’s exact testing was performed for categorical comparisons. The threshold for p-value significance was <0.05. Total symptom burden p-values were presented unadjusted as this was a summative primary outcome. P-values for individual symptoms were adjusted using the Holm-Bonferroni correction.

## Results

### General Analysis

The general analysis cohort comprised 194 patients. Within this cohort (79.9% female, median age 38 [range 18-85] years, median BMI 23.5 [range 16.2-42.1] kg/m^2^), 24.2% (n=47/194) had a delayed GET and the remaining 75.8% (n=147/194) had a normal GET. Symptom associations with GET are reported in **Table 1**. Overall, patients with a delayed GET had a significantly higher total symptom burden than patients with a normal GET (15.6 vs 8.8, p=0.006), specifically in symptoms of postprandial distress (excessive fullness (3.9 vs. 2.0, p=0.004), early satiation (6.0 vs. 1.0, p=0.015)). While increases in median symptom severity were noted across other symptoms in association with delayed gastric emptying, these did not reach statistical significance.

**TABLE 1.** Median scores for overall symptoms in delayed and normal gastric emptying groups, showing significant associations for total symptom burden, excessive fullness and early satiation. All other associations are non-significant.

| Symptom | Delayed GE | Normal GE | p-value |
| --- | --- | --- | --- |
| **Total Symptom Burden** | **15.6** | **8.8** | **0.006** |
| **Excessive Fullness** | **3.9** | **2.0** | **0.004** |
| **Early Satiation** | **6.0** | **1.0** | **0.015** |
| Nausea | 1.7 | 0.5 | 0.170 |
| Bloating | 2.3 | 0.9 | 0.432 |
| Upper Gut Pain | 1.6 | 1.0 | 0.924 |
| Reflux | 0.0 | 0.0 | 1.000 |
| Vomiting | 0.0 | 0.0 | 1.000 |
| Heartburn | 0.0 | 0.0 | 1.000 |
| Belching | 0.0 | 0.0 | 1.000 |
| Stomach Burn | 0.0 | 0.0 | 1.000 |

Data expressed as median scores. Significance defined as $p < 0.05$ .

Additional analyses were conducted focused on the correlation between symptom burden and GET (t_1/2_ emptying time (GEBT) and % retention at 4 hours (GES)). For the GEBT dataset, there was a significant although weak correlation between total symptom burden and t_1/2_ emptying time (r_s_=0.30, p<0.001) (**Figure 1**), as well as for multiple individual symptoms including: excessive fullness (r_s_=0.28, p=0.011), nausea (r_s_=0.28, p=0.011), early satiation (r_s_=0.26, p=0.020), bloating (r_s_=0.24, p=0.041), and upper gut pain (r_s_=0.24, p=0.041). For the GES dataset, there were no correlations found, likely reflecting limited variance at the four-hour endpoint, as a significant proportion of the cohort had already achieved complete emptying (0% retention).

**FIGURE 1.**
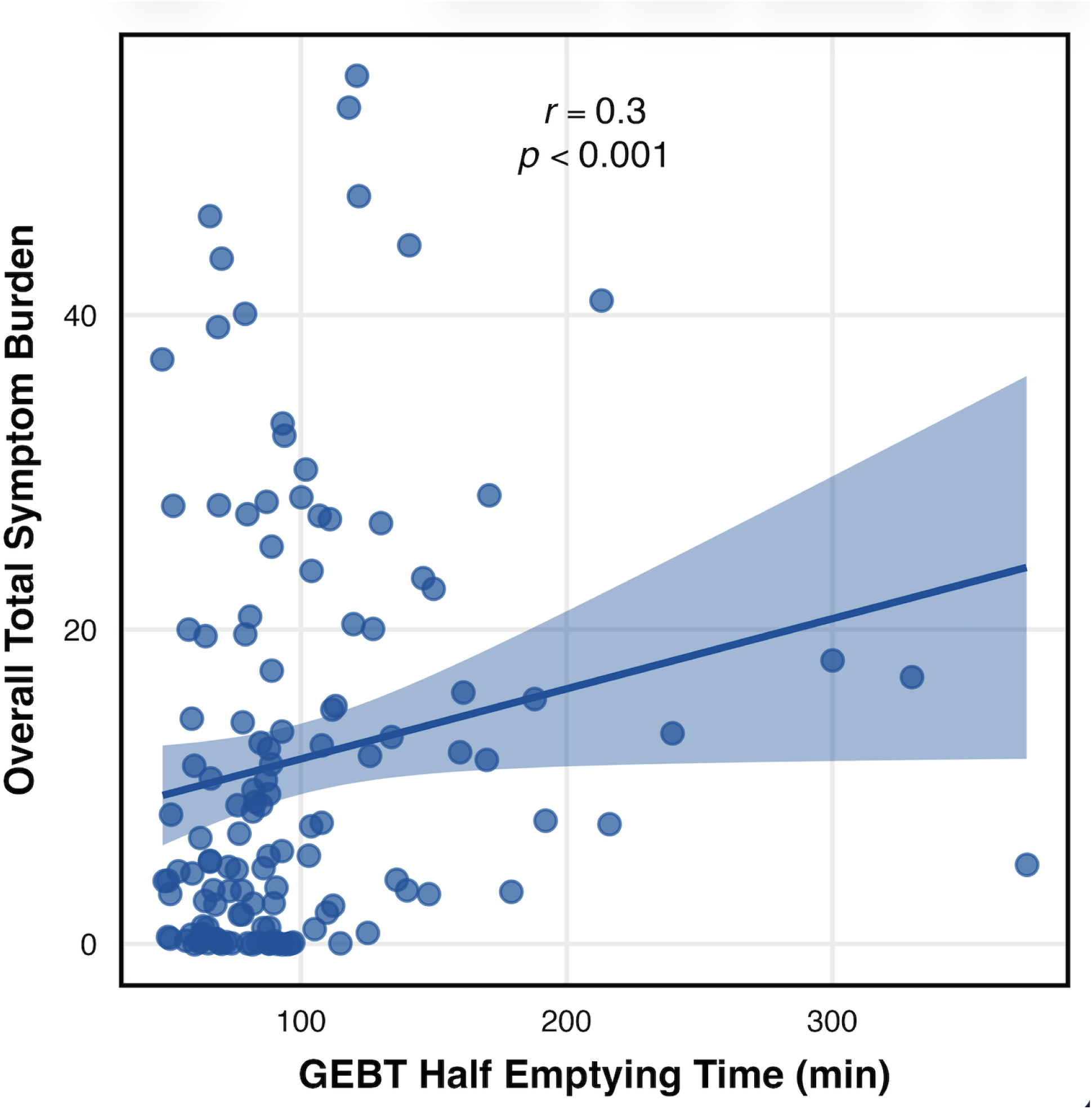
Scatter plot demonstrating a weak correlation (Spearman’s rank) between overall total symptom burden and half emptying time in the GEBT dataset (n=143). The shaded region represents the 95% confidence interval.

A further analysis of symptom associations was conducted during the intra-test time intervals, i.e. preprandial, and first, second, third, and fourth-hour postprandial periods. For total symptom burden and excessive fullness, there was a consistent and significant association between symptom severity and delayed gastric emptying, and this association was notably strongest in the first postprandial hour before progressively reducing in strength throughout the remaining postprandial test period (**Figure 2**). In addition, preprandial (baseline) symptoms were also greater in those with delayed gastric emptying (total symptom burden 7.0 vs. 2.6, p=0.009).

**FIGURE 2.**
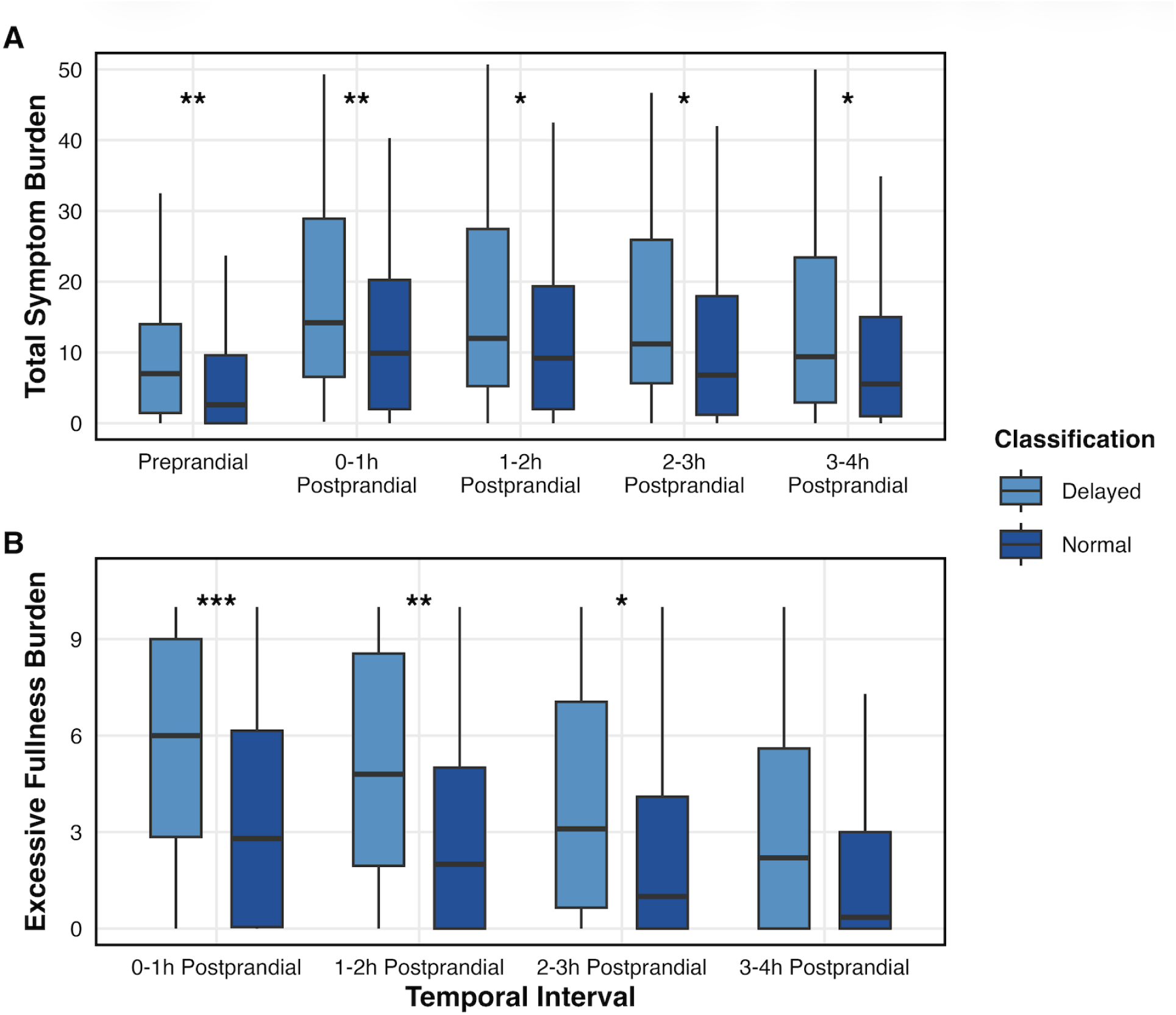
Box plots showing the temporal progressive reduction in symptom burden for total symptom burden (A) and excessive fullness (B). The excessive fullness plot does not have a preprandial measure in this study. Significance levels indicate the difference between delayed and normal gastric emptying groups: * p<0.05; **p<0.01; *** p<0.001.

### ‘Auckland Classification’ Analysis

‘Auckland Classification’ phenotyping was undertaken on 155 patients. Within this cohort (80.6% female, median age 33 [range 18-85] years, median BMI 22.9 [range 16.7-35.0] kg/m^2^), 21.9% (n=34/155) had a delayed GET. Proportions of delayed vs. normal GET varied by phenotype: ‘Spectral Abnormal’ 24.3% (n=9/37), ‘Sensorimotor’ 29.7% (n=11/37), ‘Continuous’ 28.1% (n=9/32), and ‘N/A’ 12.7% (n=8/63). There was a significant association between being classified by any ‘Auckland Classification’ phenotype and having delayed GET (OR 2.58, 95%CI 1.05 to 7.03; p=0.034). However, between the ‘Auckland Classification’ subgroups, there were no further specific associations with delayed gastric emptying (p=0.853), i.e. no significant association between phenotype and proportion of patients with delayed GET.

Symptom associations with GET varied by ‘Auckland Classification’ phenotype (**Figures 3 and 4**). Patients with a ‘Sensorimotor’ phenotype (n=37) showed symptom associations with GET, specifically for nausea (4.9 with delayed emptying vs. 1.0 with normal emptying, p=0.027). This relationship demonstrated a moderately strong linear correlation in the GEBT cohort (r_s_=0.59, p=0.009) (**Figure 5**). Other symptoms showed similar trends within the ‘Sensorimotor’ phenotype, as illustrated by an increased overall total symptom burden (23.3 vs. 8.4, p=0.135; correlation r_s_=0.27, p=0.189), however, these did not reach statistical significance within the study sample size. By contrast, patients with a ‘Continuous’ phenotype (n=32) did not show any associations between symptoms and gastric emptying for either nausea (5.6 vs. 5.6, p=1.000; correlation r_s_=-0.11, p=0.749) or for total symptom burden (28.5 vs. 28.5, p=0.834; correlation r_s_=0.14, p=0.689). ‘Spectral Abnormal’ (n=37) phenotypes did not show any significant associations between symptoms and delayed GET, with associations for total symptom burden as follows: ‘Dysrhythmic’ (n=9) (6.8 vs. 19.5, p=0.381), ‘High Frequency’ (n=10) (18.0 vs. 5.6, p=1) and ‘Low Meal Response’ (n=18) (11.7 vs. 3.6, p=0.722). Furthermore, the unclassified (‘NA’) category (n=63) did not show any significant associations, i.e. total symptom burden (11.3 vs. 5.0, p=0.230).

**FIGURE 3.**
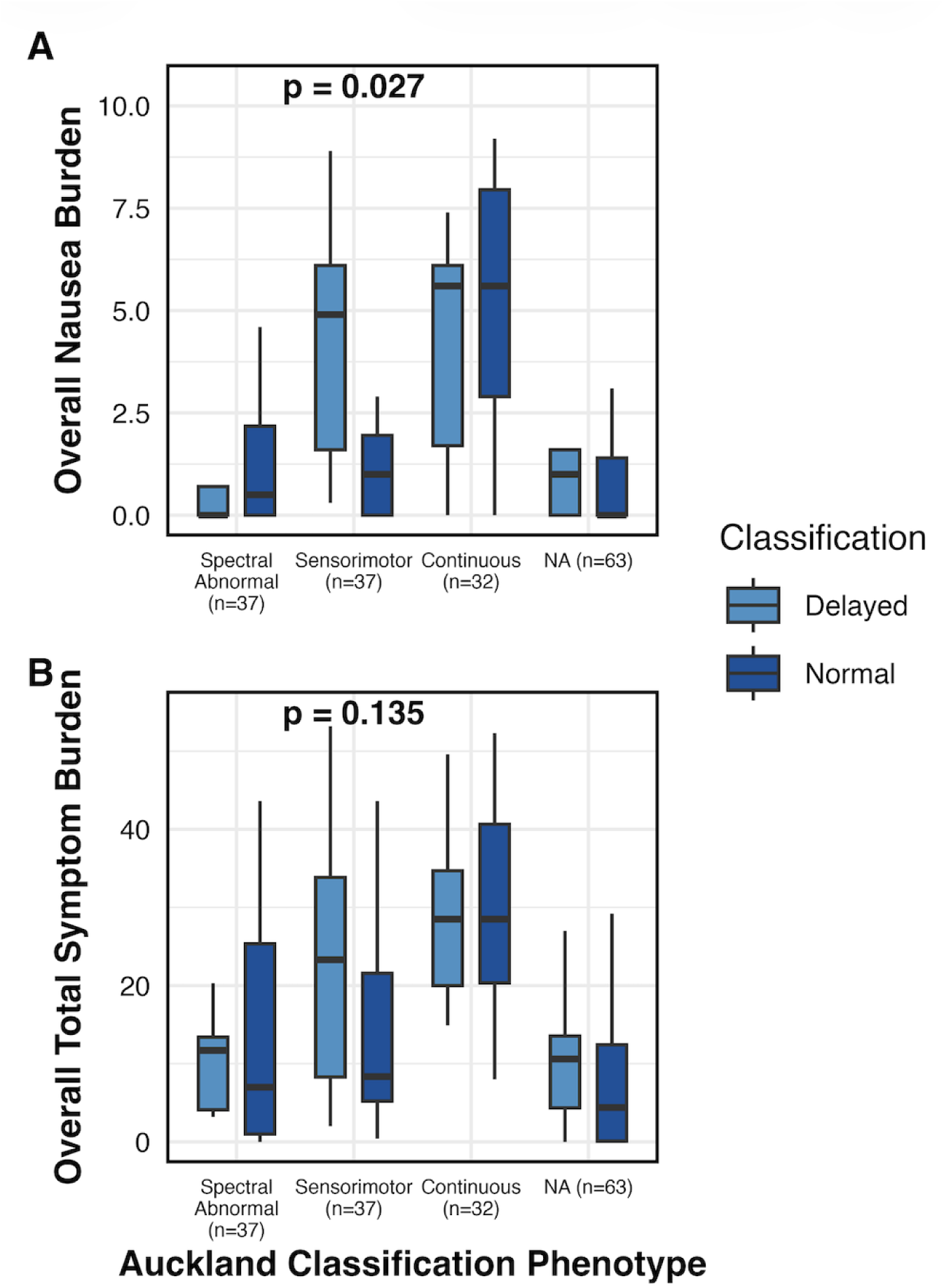
Box plots depicting median, interquartile range, and range, demonstrating associations between gastric emptying and nausea symptoms (A) and total symptom burden (B) in a population subgrouped by the ‘Auckland Classification’ scheme. Symptom associations are exclusive to the ‘Sensorimotor’ phenotype, and absent in the ‘Continuous’ phenotype and ‘Spectral Abnormal’ phenotypes. ‘Spectral Abnormal’ phenotypes include ‘Dysrhythmic’, ‘High Frequency’, and ‘Low Meal Response’.

**FIGURE 4.**
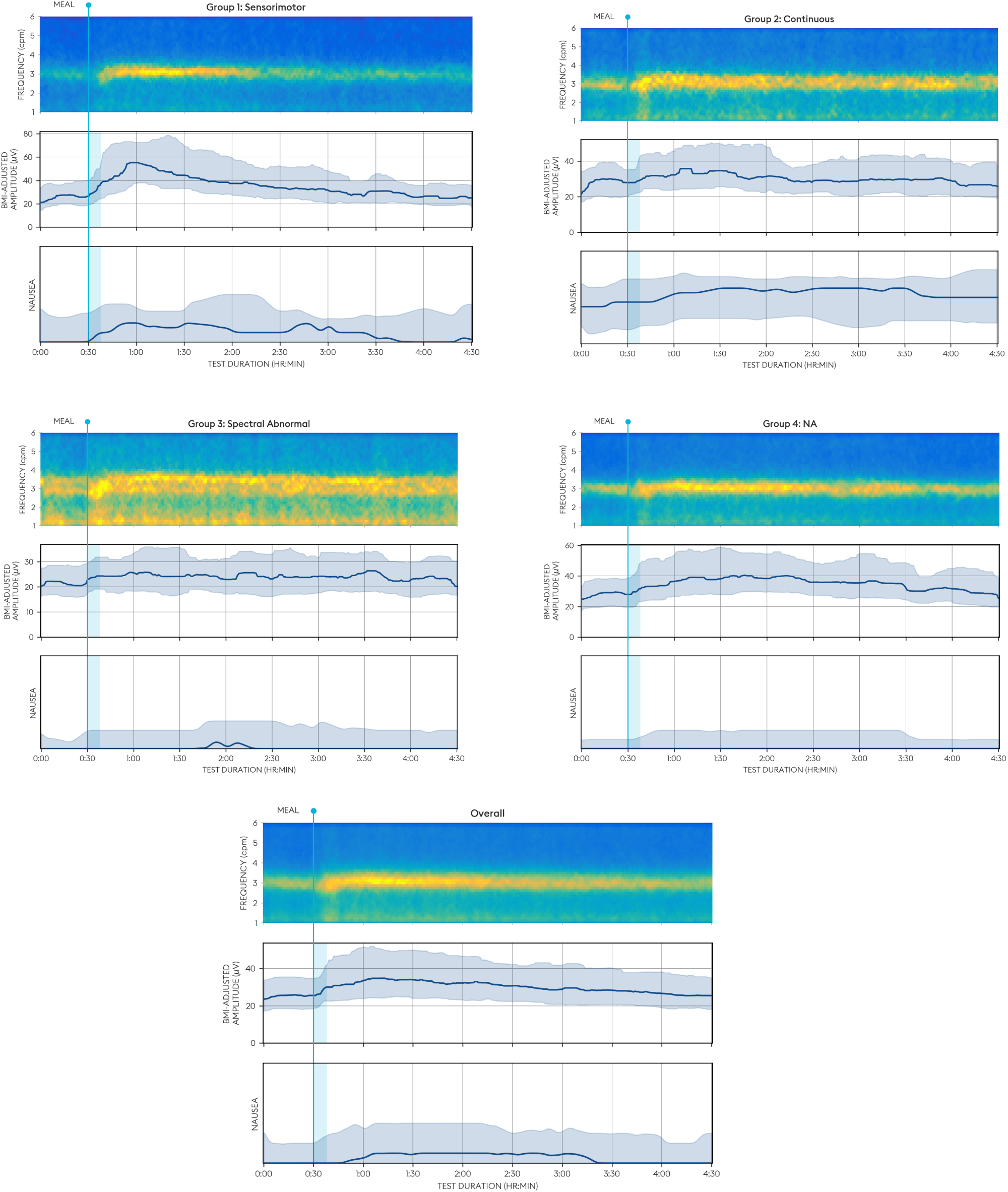
Plots showing average gastric slow wave frequency, BMI-adjusted amplitude, and nausea burdens for ‘Sensorimotor’, ‘Continuous’, ‘Spectral Abnormal’, ‘NA’ and unstratified ‘Overall’ groups.

**FIGURE 5.**
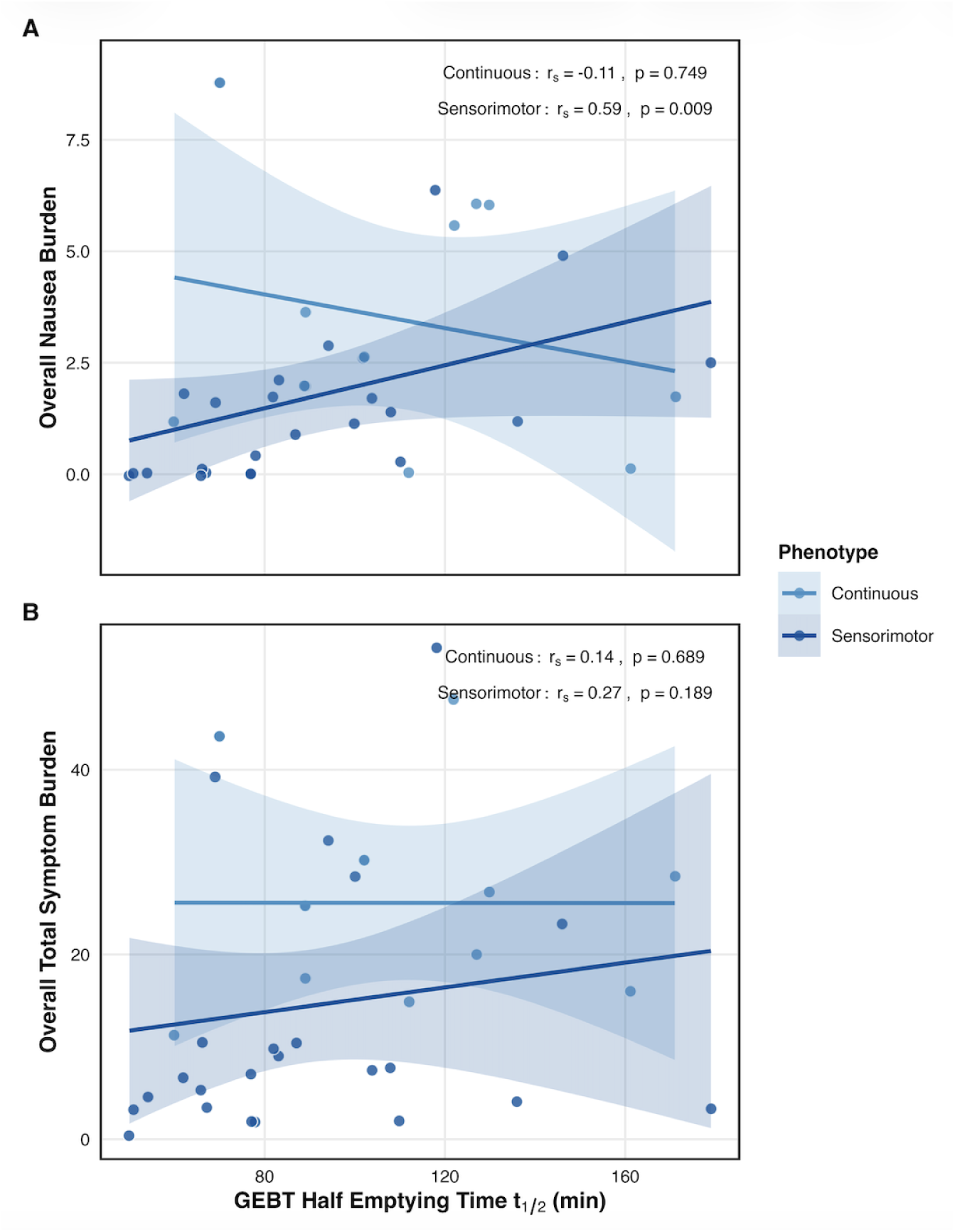
Scatter plots demonstrating a moderate correlation (Spearman’s rank) between overall nausea and half emptying time and a weak correlation (Spearman’s rank) between overall total symptom burden and half emptying time, exclusive to patients with a ‘Sensorimotor’ phenotype (n=37), and which are absent in patients with a ‘Continuous’ phenotype (n=32). The shaded regions represent the respective 95% confidence intervals.

## Discussion

This study aimed to evaluate the association between symptoms and gastric emptying in patients with chronic gastroduodenal symptoms, using time-of-test symptom analysis and subgroup profiling. The results showed a clear albeit weak overall association between symptoms and gastric emptying, most prominently for postprandial distress. However, these associations were not uniform across all patients, but were found to be dependent on specific underlying phenotypes. When our subjects were stratified by the ‘Auckland Classification’, symptom associations were exclusive to the ‘Sensorimotor’ phenotype, whereas patients with the ‘Continuous’ symptom phenotype showed almost identical symptom burdens regardless of gastric emptying. These results confirm our hypothesis that symptom associations differ according to mechanistic phenotypes within the gastroparesis and functional dyspepsia spectrum. We infer that the clinical utility of targeting gastric emptying may be highly relevant for specific subgroups while potentially negligible for others within the spectrum.

There are inconsistent findings in the literature with regards to the association between symptoms and gastric emptying. Whilst some studies cite minimal or no association, and thus question the clinical utility of gastric emptying testing and emptying-targeting interventions ^6,7,26^, our findings align with systematic review evidence that a weak association exists ^10^. We attribute our findings, in part, to the improved accuracy of time-of-test symptom profiling using a validated App. This method decreases recall bias compared to recall questionnaires ^27,28^, includes pictograms for optimal patient symptom identification ^29^, while also enabling the analysis of symptom associations throughout intra-test intervals ^16^.

In addition, this approach allowed us to demonstrate that while gastric emptying is assessed at the four-hour postprandial mark, the symptom associations with delayed gastric emptying are strikingly more prominent in the first postprandial hour. We interpret this finding as indicating that delayed emptying is unlikely to be the direct driver of symptoms, but rather serves as an indirect indicator of a more global gastric dysfunction that is active immediately upon eating. The nature of this dysfunction was not addressed in the present study, but could relate to factors such as impaired accommodation, altered intra-gastric meal distribution, and impaired postprandial antral activity, which are well known contributors to gastroduodenal symptoms ^30–32^. Furthermore, the preprandial baseline total symptom burden was already greater in those with delayed gastric emptying. As these tests were performed after a 6-8 hour fast, the presence of elevated baseline symptoms suggests that the underlying pathophysiology is not solely meal-dependent, but may reflect a chronic state of sensory or immune activation ^6,33^.

Furthermore, our study refines the association between symptoms and gastric emptying by demonstrating that this association appears to be predominantly driven by a specific subset of patients rather than a common feature of the entire disorder population. We observed that patients with a ‘Sensorimotor’ phenotype demonstrated a significant correlation between delayed gastric emptying and nausea, with an overall trend emerging for total symptom burden. The ‘Sensorimotor’ phenotype is defined in the ‘Auckland Classification’ by a high correlation between gastric electrical activity (amplitude) and symptoms ^24^. The mechanisms for this association were not assessed in the present study, but we hypothesize that the nausea symptoms could be related to a relative state of gastric hypersensitivity, where the mechanical distension associated with increased wall tension triggers an exaggerated sensory response in the vagal afferents ^34–37^. This group therefore presents a rational target population for pharmacological or interventional trials addressing gastric emptying, tone and sensitivity.

By contrast, the ‘Continuous’ phenotype showed no association with gastric emptying, with almost identical symptom burdens regardless of gastric emptying. This is consistent with emerging theory that patients with continuous symptoms suffer from a higher burden of central sensitization, as opposed to peripheral motility defects, consistent with the high psychological comorbidity burden repeatedly seen in this group ^23^. Thus, these patients’ symptoms may be hypothesized to respond less favourably towards interventions that target gastric emptying, such as prokinetics or pyloric interventions (e.g. G-POEM) ^38^. Furthermore, we argue that the distinction between ‘Sensorimotor’ and ‘Continuous’ phenotypes underscores the principle that both gastroparesis and functional dyspepsia are heterogeneous disorders that cannot be adequately managed using gastric emptying status alone ^15^. The addition of BSGM can thus facilitate deeper phenotyping insights with potential for targeted treatments, potentially alleviating some of the gaps in clinical management of these disorders ^39^. Future research is now needed to test and validate this hypothesis.

Several limitations of this study are noted. First, this pooled analysis utilized data from two different modalities, both GEBT and GES. While both modalities are validated, discrepancies in test meals and additional confounding factors may have introduced some degree of variability. However, it can be noted that test meals were essentially calorie- and macronutrient-equivalent. In addition to this, to mitigate modality differences between GEBT and GES, we utilized standardized, modality-specific cut-offs for delayed emptying. Second, the use of a binary definition for delayed and normal GET is inherently arbitrary and does not necessarily capture the continuous nature of biological function. This was accounted for by also analyzing correlations between symptoms and raw continuous data (half emptying time (GEBT) and percentage retention at 4 hours (GES)). However, the overwhelming number of GES patients with 0% retention at 4-hours postprandial limited the relevance of these analyses to the GEBT cohort alone. Third, the sample sizes for specific ‘Auckland Classification’ phenotypes, particularly ‘Dysrhythmic’ and ‘High Frequency’, were relatively small in these cohorts of community patients referred for emptying testing, at n=9 and n=10 respectively, thus limiting the statistical power to detect subtle associations within these further subgroups. This provides opportunities for future studies to recruit more patients and conduct specific analyses for these subgroups. Fourth, and linked to the aforementioned point, some patients were unclassified and assigned to the ‘NA’ phenotype (n=63, 37.3%). Although delayed emptying was found to be uncommon in this group, it is possible that they suffer from mechanisms that are not directly detected by either gastric emptying testing or BSGM, such as disordered gastric accommodation. Further research is ongoing to more fully characterise this group of patients.

In conclusion, this study demonstrates a weak but clear relationship between symptoms and gastric emptying when optimal time-of-test symptom profiling is applied. Crucially, this study also moves beyond a ‘one-size-fits-all’ approach to gastroparesis by utilizing the novel ‘Auckland Classification’ scheme to identify that these symptoms associations are restricted to the ‘Sensorimotor’ phenotype and are definitively absent in ‘Continuous’ patients. These findings suggest that future clinical management should prioritize physiological phenotyping to identify those patients who are most likely to benefit from gastric emptying testing and emptying-targeted therapies, as opposed to other care modalities.

## Data Availability

Data used for analysis will be made available upon reasonable request, conditional on ethical approvals.

