## Supplementary Figures 1-3 for "Symptom Associations with Delayed Gastric Emptying Vary by Disease Phenotype"

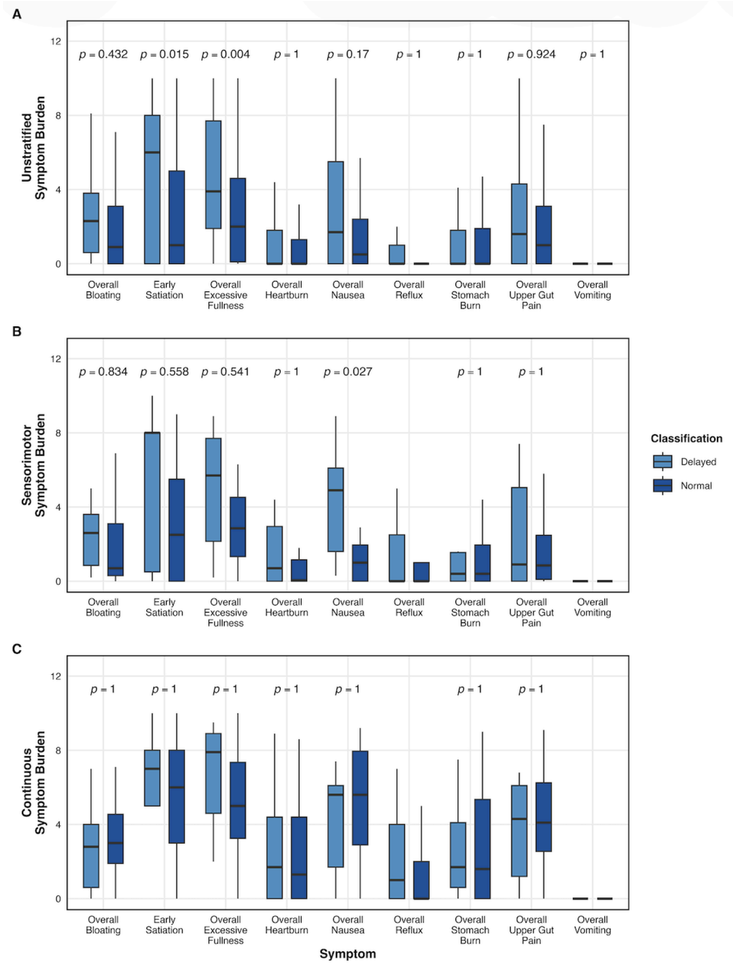


**SUPPLEMENTARY FIGURE 1** | Comparison between delayed and normal gastric emptying for individual symptoms in the overall unstratified population (A), in a subgroup of individuals with a 'Sensorimotor' phenotype (B), and in a subgroup of individuals with a 'Continuous' phenotype (C). In the overall unstratified population, there are significant associations for early satiation and excessive fullness. Following 'Auckland Classification' phenotyping, in the 'Sensorimotor' subgroup, there is a significant association exclusive for nausea, whilst in the 'Continuous' subgroup, there are no significant associations.

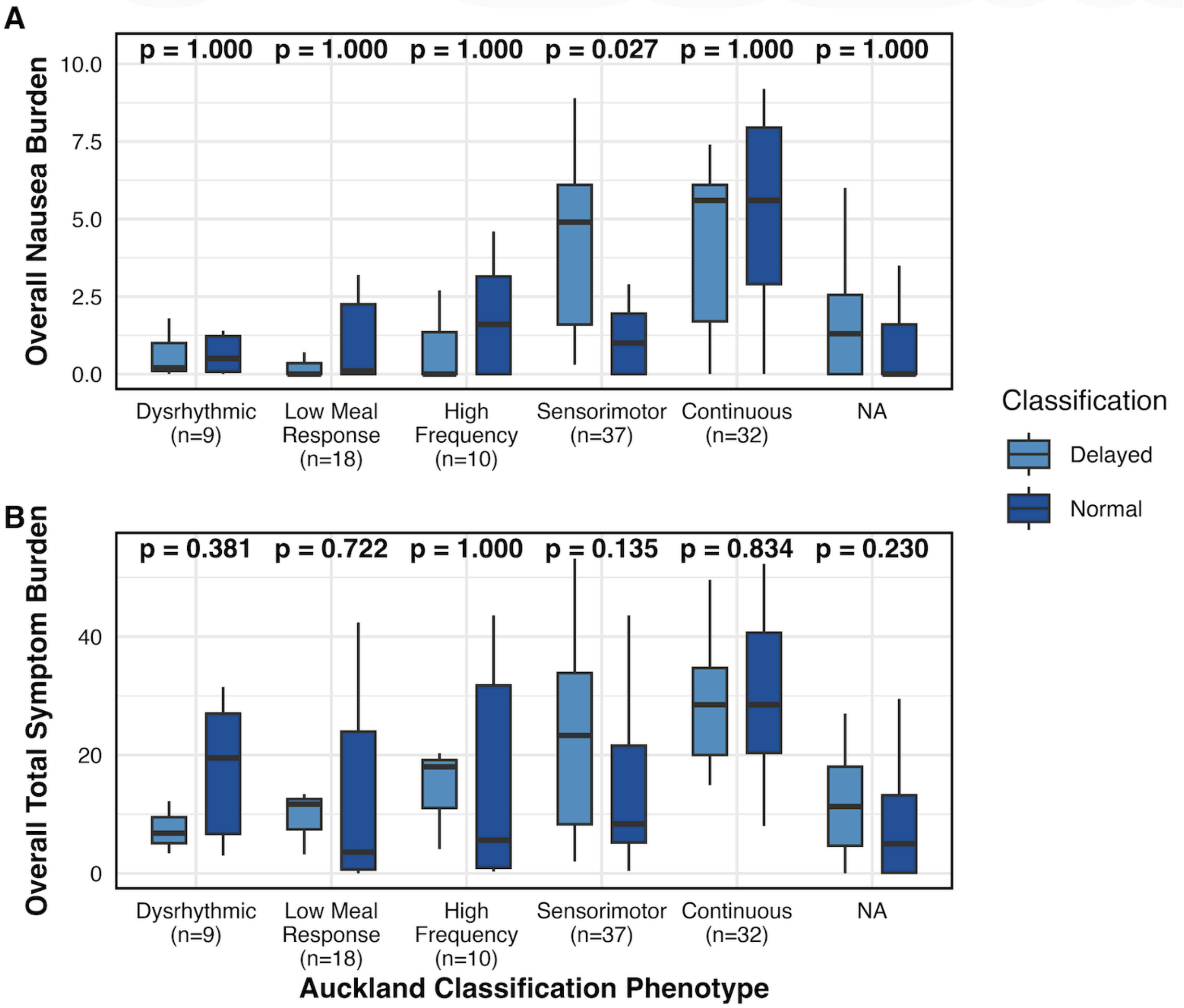


**SUPPLEMENTARY FIGURE 2** | Box plots depicting median, interquartile range, and range, demonstrating associations between nausea symptoms (A) and total symptom burden (B) with gastric emptying in a population subgrouped by the ‘Auckland Classification’ scheme. Symptom associations are exclusive to the ‘Sensorimotor’ phenotype, and absent in the ‘Continuous’ phenotype. Symptom associations are also not found in the ‘Dysrhythmic’, ‘High Frequency’, and ‘Low Meal Response’ phenotypes.

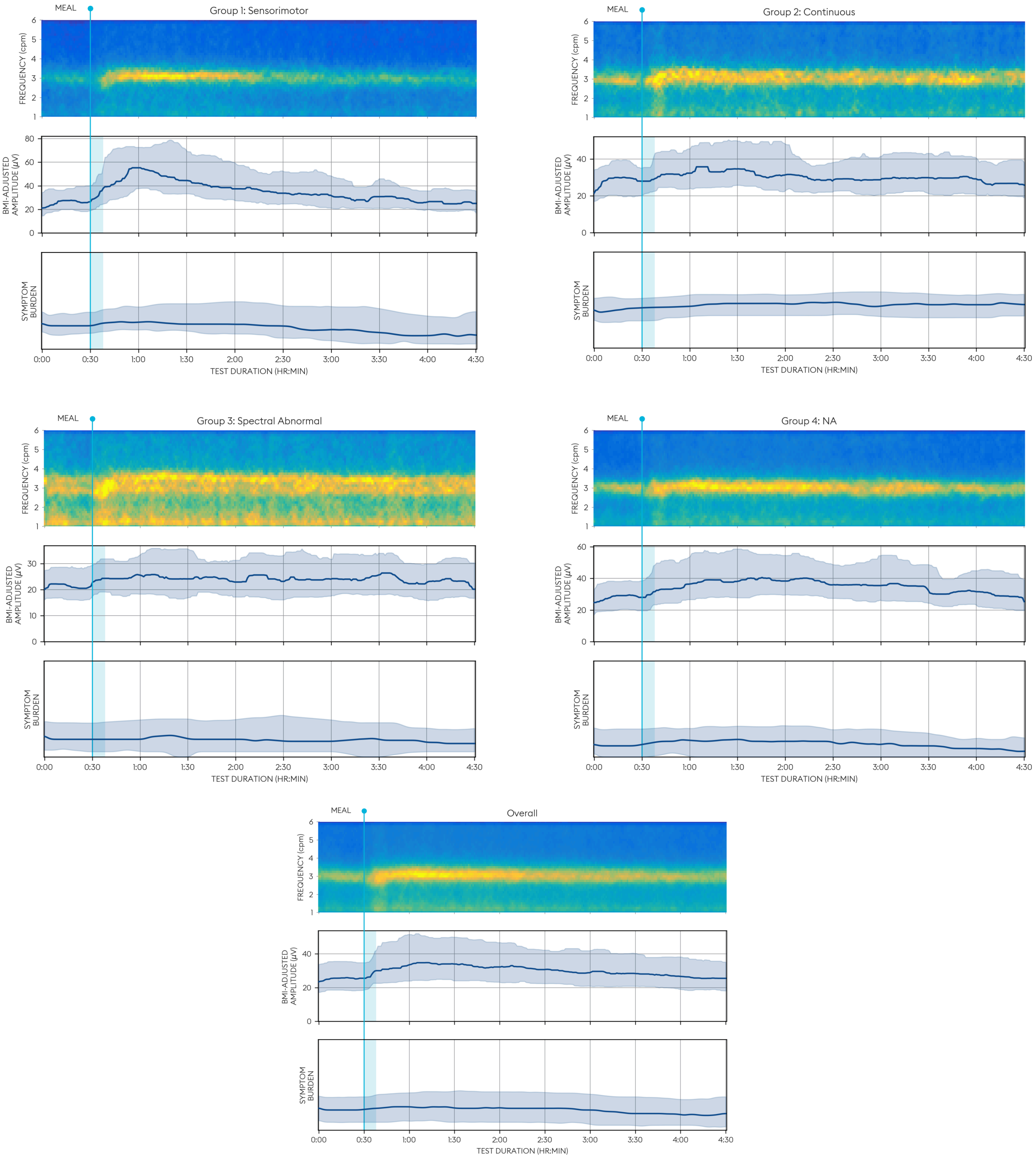

MEAL
Group 1: Sensorimotor
FREQUENCY (cpm)
6
5
4
3
2
1
BMI-ADJUSTED AMPLITUDE (µV)
80
60
40
20
0
SYMPTOM BURDEN
0:00
0:30
1:00
1:30
2:00
2:30
3:00
3:30
4:00
4:30
TEST DURATION (HR:MIN)
MEAL
Group 2: Continuous
FREQUENCY (cpm)
BMI-ADJUSTED AMPLITUDE (µV)
40
20
0
SYMPTOM BURDEN
TEST DURATION (HR:MIN)
MEAL
Group 3: Spectral Abnormal
FREQUENCY (cpm)
BMI-ADJUSTED AMPLITUDE (µV)
30
20
10
0
SYMPTOM BURDEN
TEST DURATION (HR:MIN)
MEAL
Group 4: NA
FREQUENCY (cpm)
BMI-ADJUSTED AMPLITUDE (µV)
60
40
20
0
SYMPTOM BURDEN
TEST DURATION (HR:MIN)
MEAL
Overall
FREQUENCY (cpm)
BMI-ADJUSTED AMPLITUDE (µV)
40
20
0
SYMPTOM BURDEN
TEST DURATION (HR:MIN)

**SUPPLEMENTARY FIGURE 3** | Plots showing average gastric slow wave frequency, BMI-adjusted amplitude, and total symptom burdens for ‘Sensorimotor’, ‘Continuous’, ‘Spectral Abnormal’, ‘NA’ and unstratified ‘Overall’ groups.
